# Multiparametric CT Phenotypes Identify Risks of Early Neurological Deterioration and 90-Day Disability in Minor Stroke

**DOI:** 10.64898/2026.09.21.26363615

**Authors:** Francisco Purroy, Eugènia Saureu-Rufach, Laura Pérez-Girona, Albert Freixa-Cruz, Alberto Martínez-Leza, Cristina Pereira, Ares Peguera, Sara Salvany, Raquel Mitjana, Gloria Arqué, Yhovany Gallego, Mikel Vicente-Pascual, Anna Fernández-Bernal, Gerard Mauri-Capdevila

## Abstract

**Background:** Minor ischemic stroke is heterogeneous, and low NIHSS does not reliably identify patients at risk of deterioration or disability. We investigated whether integrating CT perfusion and angiographic vessel status identifies prognostically distinct phenotypes in a consecutive stroke-code cohort.

**Methods:** We analyzed consecutive adults with ischemic stroke, baseline NIHSS 0-5, prestroke modified Rankin Scale (mRS) ≤2, presentation within 24 hours of symptom onset or last known well, and emergency multiparametric CT. Patients were classified as no perfusion deficit/no vessel occlusion (PF-/VO-), perfusion deficit without visible occlusion (PF+/VO-), or perfusion deficit with vessel occlusion (PF+/VO+). The primary outcome was mRS ≥3 at 90 days; early neurological deterioration (END), defined as an NIHSS increase ≥2 points by 24 hours, was secondary. Multivariable logistic regression adjusted for prespecified covariates and additional variables associated with each outcome in univariable analysis.

**Results:** Of 2211 patients evaluated through the stroke-code pathway, 534 met inclusion criteria; 391 (73.2%) were PF-/VO-, 61 (11.4%) PF+/VO-, and 82 (15.4%) PF+/VO+. END occurred in 8.1% and mRS ≥3 at 90 days in 17.6%. END occurred in 6.6%, 4.9%, and 17.1% of PF-/VO-, PF+/VO-, and PF+/VO+ patients, respectively (P=0.004), whereas mRS ≥3 occurred in 13.3%, 34.4%, and 25.6%, respectively (P<0.001). Compared with PF-/VO-, PF+/VO+ was independently associated with END (adjusted odds ratio [aOR], 2.50 [95% CI, 1.03-6.06]), whereas PF+/VO- was independently associated with mRS ≥3 at 90 days (aOR, 3.36 [95% CI, 1.67-6.73]). Findings were consistent across definitions of disabling versus nondisabling stroke and after exclusion of lacunar stroke.

**Conclusions:** Multiparametric CT identified distinct risk phenotypes in minor ischemic stroke. Perfusion deficit without visible vessel occlusion was associated with 90-day disability, whereas combined perfusion deficit and vessel occlusion identified patients at increased risk of END. Tissue perfusion and vessel status may provide complementary information for acute risk stratification and future trial enrichment.

## Introduction

Minor ischemic stroke (MIS), commonly defined as a National Institutes of Health Stroke Scale (NIHSS) score ≤5, accounts for approximately half of acute ischemic stroke presentations.^1,2^ Despite mild initial deficits, MIS is not uniformly benign; a clinically relevant proportion of patients develop early neurological deterioration (END) or remain disabled at 90 days.^1,3–5^ These observations challenge the assumption that low NIHSS reliably reflects low biological risk.

Current treatment decisions rely heavily on whether the presenting deficit is considered disabling. Contemporary American Heart Association/American Stroke Association guidance supports intravenous thrombolysis in otherwise eligible patients with disabling deficits, whereas thrombolysis is generally not recommended for nondisabling MIS, for which antiplatelet-based medical treatment is favored.^6^ This distinction derives from the established benefit of thrombolysis in acute ischemic stroke and the absence of demonstrated superiority in trials specifically enrolling patients with nondisabling or mild stroke.^7–11^ However, the boundary between disabling and nondisabling stroke remains inherently context dependent. PRISMS, which compared alteplase with aspirin in minor nondisabling stroke, operationalized disability according to the expected effect of the deficit on activities of daily living or return to work, whereas ARAMIS, which compared dual antiplatelet therapy with alteplase, used a more structured NIHSS item-based definition.^8,9^ Consequently, there remains no universally accepted operational definition that fully captures the functional relevance of a minor neurological deficit.

Patients with mild deficits were also underrepresented in the pivotal randomized trials that established intravenous thrombolysis and endovascular thrombectomy, leaving uncertainty regarding the optimal treatment of this population.^7,12,13^ Dedicated minor-stroke trials have not resolved this uncertainty. PRISMS^8^ was terminated early and did not demonstrate benefit of alteplase over aspirin; ARAMIS^9^ showed noninferiority of dual antiplatelet therapy to alteplase; PUMICE^10^ found no superiority of prourokinase over standard care; and TEMPO-2^11^ found no benefit of tenecteplase in patients with minor stroke selected by intracranial occlusion or focal perfusion abnormality. These neutral results may in part reflect the biological heterogeneity of MIS and the limitations of selection based predominantly on clinical severity or a single imaging feature.

Multiparametric computed tomography offers an objective, tissue-based complement to clinical assessment. CT angiography (CTA) identifies intracranial vessel occlusion, whereas CT perfusion (CTP) characterizes the extent and severity of cerebral hypoperfusion.^14^ The value of advanced imaging for treatment selection is well established in extended or uncertain time windows for reperfusion, including trials based on tissue viability and perfusion mismatch.^15–18^ In minor stroke, however, most available prognostic evidence derives from selected populations with known large-vessel occlusion (LVO), anterior-circulation stroke, or specific reperfusion strategies. In these settings, occlusion characteristics and perfusion abnormalities have been associated with END and poorer outcomes.^3,19,20^

The prognostic significance of perfusion abnormalities in an unselected stroke-code MIS population therefore remains incompletely defined, particularly in patients without a visible intracranial occlusion. Vessel status and tissue perfusion are related but not interchangeable: measurable hypoperfusion may persist despite the absence of an identifiable occlusion, while the hemodynamic consequences of a visible occlusion may vary according to collateral circulation, recanalization, and tissue vulnerability. We therefore evaluated consecutive patients with MIS undergoing emergency multiparametric CT to determine whether the combined assessment of perfusion deficit and vessel occlusion identifies distinct phenotypes associated with END and 90-day functional outcome.

## Methods

### Study Design and Participants

We conducted a single-center observational cohort study including consecutive patients evaluated through the institutional stroke-code pathway at a tertiary university hospital between May 2021 and August 2025. Adults were eligible if the final diagnosis was acute ischemic stroke, baseline NIHSS was 0-5, prestroke modified Rankin Scale (mRS) was ≤2, symptom onset or last known well was within 24 hours, and emergency multiparametric CT was performed as part of the acute evaluation. Patients were excluded from the analytic cohort for prestroke mRS >2, absence of CTP, or technically invalid image reconstruction. Reporting followed STROBE recommendations.^21^

Baseline demographic characteristics, vascular risk factors, previous ischemic stroke, witnessed onset, baseline NIHSS, and time from symptom onset or last known well to imaging were recorded prospectively within the stroke-code workflow. Baseline NIHSS was additionally categorized as 0, 1-2, or 3-5. The presenting clinical syndrome was classified according to the Oxfordshire Community Stroke Project classification as partial anterior circulation infarction, posterior circulation infarction, or lacunar infarction.^22^ Stroke etiology was adjudicated after completion of the diagnostic work-up according to the Trial of ORG 10172 in Acute Stroke Treatment (TOAST) classification.^23^

Because reperfusion decisions in MIS often depend on whether symptoms are considered disabling, two operational definitions were applied. The primary definition followed the PRISMS framework: deficits were considered disabling when, if unchanged, they would be expected to prevent basic activities of daily living or return to work or usual activities.^8^ As a sensitivity framework, nondisabling MIS was also classified according to the item-based ARAMIS criteria.^9^

All examinations were performed on a 128-detector CT scanner (SOMATOM Definition Edge VB20; Siemens Healthineers). The emergency multiparametric CT protocol comprised noncontrast CT (NCCT), CT angiography (CTA), and CT perfusion (CTP). NCCT was acquired at 120 kV and 320 mAs with 2-mm slice thickness and 1-mm reconstructed images. CTP provided 84-mm craniocaudal brain coverage and was acquired at 70 kV and 200 mAs using 40 acquisitions over 59.23 seconds. A total of 50 mL of iodinated contrast was administered at 5.5 mL/s followed by a 40-mL saline flush, with an 8-second acquisition delay. CTA extended from below the aortic arch to the cranial vertex and was acquired at 100 kV and 175 mAs after administration of 50 mL of iodinated contrast (Xenetix 350 mg iodine/mL) at 5.5 mL/s, followed by a 40-mL saline flush; a 3-second triggering delay was used. CTA images were reconstructed at 0.75-mm slice thickness with a 0.4-mm increment. Automatic dose modulation was used for NCCT and CTA, whereas CTP was acquired with fixed tube settings.

CTP data were automatically processed using RAPID software (version 5.8; iSchemaView, Inc., Menlo Park, CA, USA), generating ischemic-core volume (relative cerebral blood flow <30%), hypoperfused-tissue volume (Tmax >6 seconds), and mismatch volume. A perfusion deficit (PF+) was defined as any measurable tissue volume with Tmax >6 seconds. CTA was used to determine intracranial vessel occlusion and its location. LVO comprised intracranial internal carotid artery, tandem occlusion, M1, proximal M2, and basilar artery occlusion; medium-vessel occlusion (MeVO) comprised distal M2, M3, anterior cerebral artery, and posterior cerebral artery occlusions. Occlusion status and the quality and validity of RAPID-derived perfusion data were independently reviewed by two neuroradiologists. Perfusion abnormalities attributable to imaging artifacts or technical limitations were excluded before CT phenotype assignment. Discordant cases were jointly reviewed with the corresponding author until consensus was reached.

Patients were classified a priori into three multiparametric CT phenotypes: PF-/VO-, PF+/VO-, and PF+/VO+. No PF-/VO+ phenotype was observed. Acute reperfusion treatment was categorized as no reperfusion, intravenous thrombolysis alone, or endovascular treatment; for regression analyses, receipt of any reperfusion therapy was analyzed as a binary variable.

The primary outcome was unfavorable functional outcome at 90 days, defined as mRS ≥3. The 90-day assessment was performed by trained personnel blinded to the results of the emergency multiparametric CT. When a dedicated 90-day assessment could not be obtained, the mRS documented at the most recent available clinical follow-up was used; patients who died were assigned mRS 6. The secondary outcome was END, prespecified as an increase of ≥2 NIHSS points by 24 hours compared with baseline, reflecting the concept of early stroke progression evaluated in contemporary minor-stroke trials including TEMPO-2 and ARAMIS.^9,11^

This study was conducted as a substudy of the Omic is Brain project and was approved by the local research ethics committee, the Comitè d’Ètica i Investigació Clínica de l’Hospital Universitari Arnau de Vilanova de Lleida (CEIC ID: 2343). All participants or their legal representatives received information about the study and provided written informed consent before inclusion.

### Statistical Analysis

Continuous variables are reported as mean (SD) or median (interquartile range [IQR]) according to distribution, and categorical variables as number (percentage). Between-group comparisons used Student t test or one-way ANOVA for normally distributed continuous variables, Mann-Whitney U or Kruskal-Wallis tests for nonnormally distributed variables, and chi-square or Fisher exact tests for categorical variables as appropriate. All tests were 2-sided, with P<0.05 considered statistically significant.

Multivariable binary logistic regression analyses were performed separately for the primary outcome (90-day mRS ≥3) and the secondary outcome (END). Age, prestroke mRS category, baseline NIHSS category, multiparametric CT phenotype, receipt of any reperfusion therapy, and PRISMS-defined nondisabling MIS were included a priori in the models on the basis of their clinical relevance. Additional covariates associated with the corresponding outcome in univariable analyses at P<0.05 were also included. The same modeling strategy was applied to both outcomes. All covariates were entered simultaneously using the enter method. PF-/VO-, NIHSS 0, and prestroke mRS 0 were used as reference categories. Adjusted odds ratios (aORs) with 95% CIs were reported.

Two prespecified sensitivity strategies were applied. First, the multivariable models were repeated after replacing the PRISMS definition with the ARAMIS definition of nondisabling MIS while retaining the remaining covariates. Second, because all TOAST small-vessel (lacunar) strokes were classified as PF-/VO-, univariable and multivariable analyses were repeated after exclusion of lacunar etiology. In the nonlacunar cohort, adjusted models were estimated using both PRISMS and ARAMIS definitions. Analyses were performed with IBM SPSS Statistics, version 29 (IBM Corp).

### Data Availability

Requests for access to the data reported in this article will be considered by the corresponding author on a reasonable basis.

## Results

### Study Population and Multiparametric CT Phenotypes

Between May 2021 and August 2025, 2211 consecutive patients evaluated through the stroke-code pathway were assessed for eligibility. Of these, 1296 (58.6%) were diagnosed with ischemic stroke, whereas 639 (28.9%) were classified as stroke mimics, 226 (10.2%) as intracerebral hemorrhage, 21 (0.9%) as subarachnoid hemorrhage, 28 (1.3%) as subdural hemorrhage, and 1 (<0.1%) as cerebral venous thrombosis. Among patients with ischemic stroke, 632 (48.8%) had a baseline NIHSS score >5 and were excluded, leaving 664 (51.2%) patients fulfilling the clinical definition of MIS. Of these, 107 (16.1%) were excluded because of prestroke mRS >2, 21 (3.2%) because CTP was not performed, and 2 (0.3%) because of image reconstruction errors. The final study population therefore comprised 534 patients (80.4% of the MIS cohort; Figure 1).

**Figure 1.**
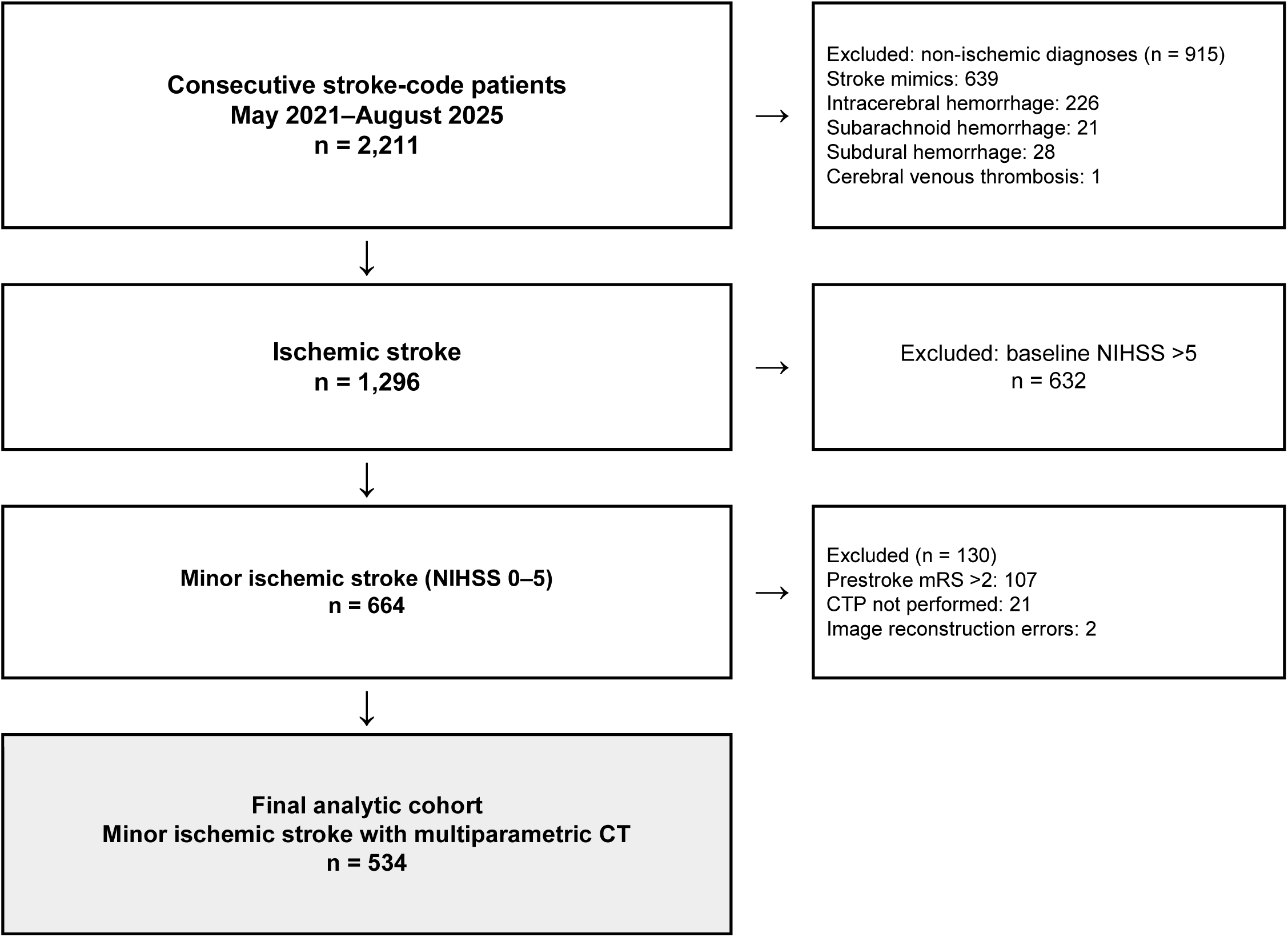
Study flowchart. Flow of consecutive stroke-code patients from initial evaluation to the final analytic cohort of 534 patients with minor ischemic stroke undergoing multiparametric CT. CTP indicates CT perfusion; mRS, modified Rankin Scale; NIHSS, National Institutes of Health Stroke Scale.

Mean age was 68.3±13.5 years, and 352 patients (65.9%) were men. Three multiparametric CT phenotypes were identified: PF-/VO- in 391 patients (73.2%), PF+/VO- in 61 (11.4%), and PF+/VO+ in 82 (15.4%). Among PF+/VO+ patients, 53 (64.6%) had LVO and 29 (35.4%) had MeVO. Clinical characteristics differed across CT phenotypes, with greater baseline neurological severity and a lower proportion of nondisabling presentations in PF+/VO+, according to both PRISMS and ARAMIS criteria. Several vascular risk factors were also unevenly distributed across phenotypes (Table 1).

**Table 1.**
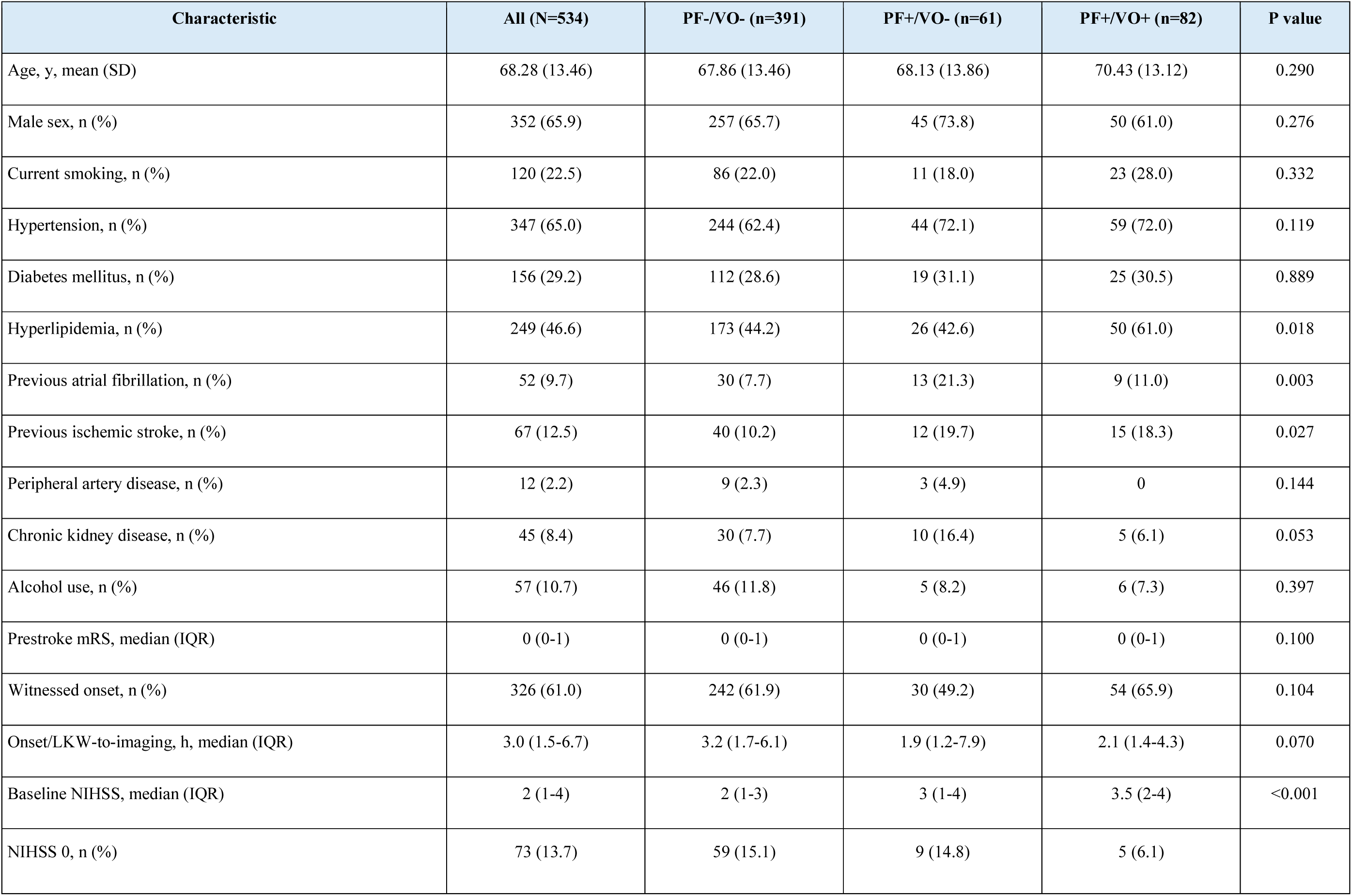
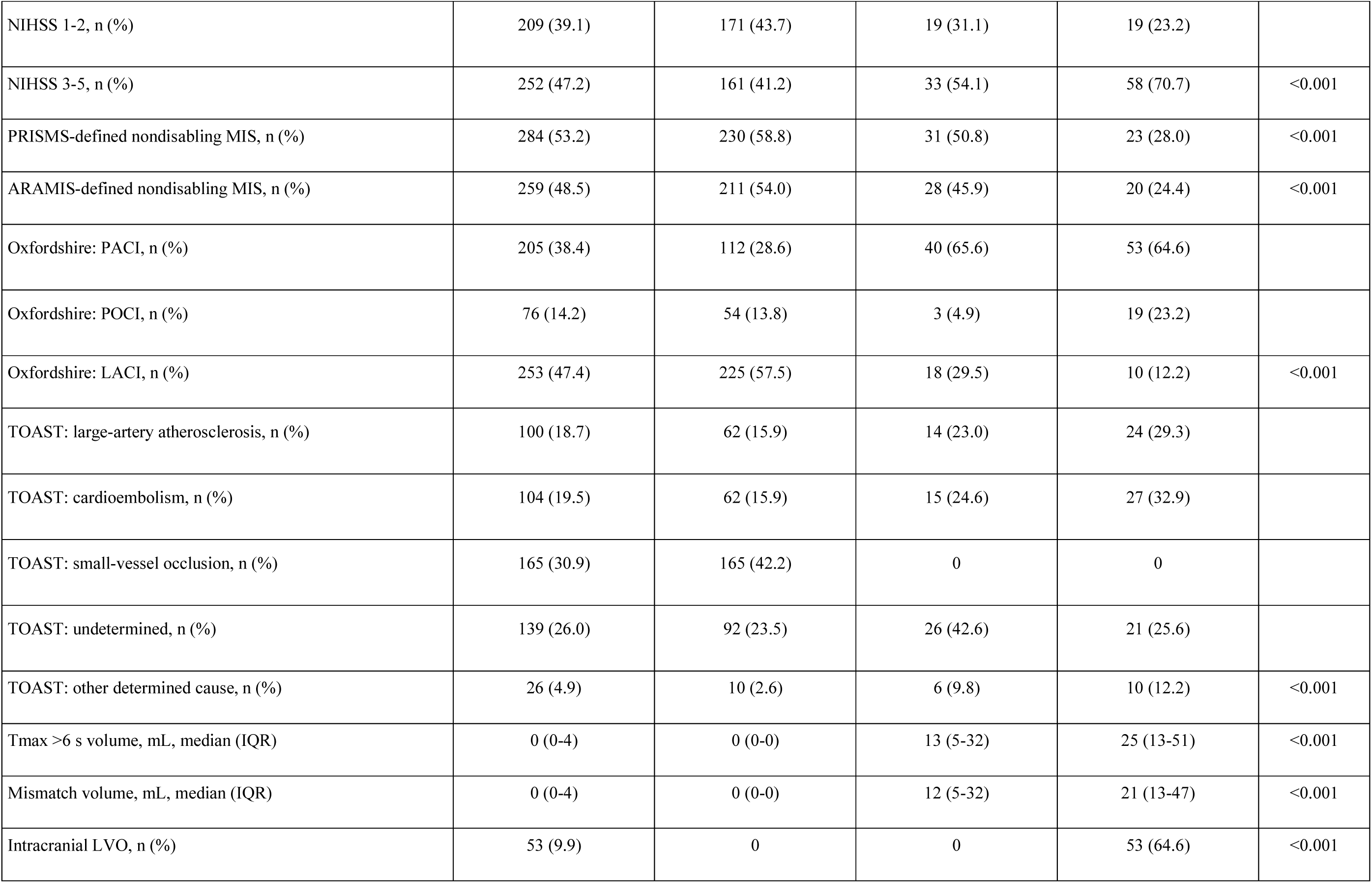
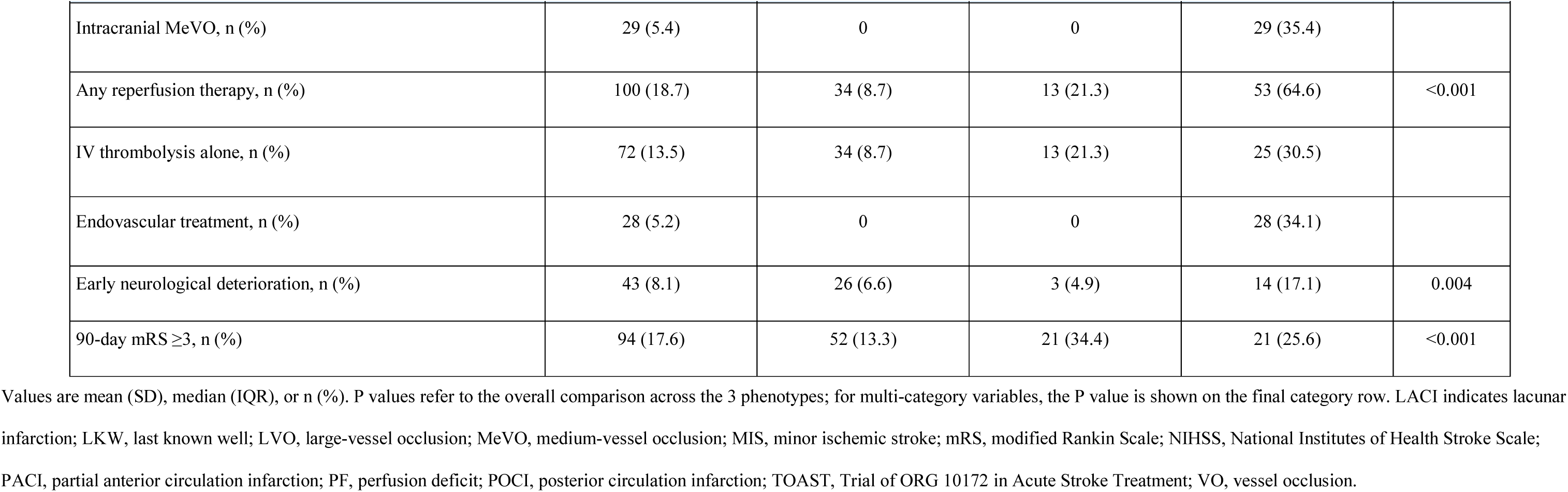
Clinical, Etiologic, Imaging, and Treatment Characteristics According to Multiparametric CT Phenotype.

| Characteristic | All (N=534) | PF-/VO- (n=391) | PF+/VO- (n=61) | PF+/VO+ (n=82) | P value |
| --- | --- | --- | --- | --- | --- |
| Age, y, mean (SD) | 68.28 (13.46) | 67.86 (13.46) | 68.13 (13.86) | 70.43 (13.12) | 0.290 |
| Male sex, n (%) | 352 (65.9) | 257 (65.7) | 45 (73.8) | 50 (61.0) | 0.276 |
| Current smoking, n (%) | 120 (22.5) | 86 (22.0) | 11 (18.0) | 23 (28.0) | 0.332 |
| Hypertension, n (%) | 347 (65.0) | 244 (62.4) | 44 (72.1) | 59 (72.0) | 0.119 |
| Diabetes mellitus, n (%) | 156 (29.2) | 112 (28.6) | 19 (31.1) | 25 (30.5) | 0.889 |
| Hyperlipidemia, n (%) | 249 (46.6) | 173 (44.2) | 26 (42.6) | 50 (61.0) | 0.018 |
| Previous atrial fibrillation, n (%) | 52 (9.7) | 30 (7.7) | 13 (21.3) | 9 (11.0) | 0.003 |
| Previous ischemic stroke, n (%) | 67 (12.5) | 40 (10.2) | 12 (19.7) | 15 (18.3) | 0.027 |
| Peripheral artery disease, n (%) | 12 (2.2) | 9 (2.3) | 3 (4.9) | 0 | 0.144 |
| Chronic kidney disease, n (%) | 45 (8.4) | 30 (7.7) | 10 (16.4) | 5 (6.1) | 0.053 |
| Alcohol use, n (%) | 57 (10.7) | 46 (11.8) | 5 (8.2) | 6 (7.3) | 0.397 |
| Prestroke mRS, median (IQR) | 0 (0-1) | 0 (0-1) | 0 (0-1) | 0 (0-1) | 0.100 |
| Witnessed onset, n (%) | 326 (61.0) | 242 (61.9) | 30 (49.2) | 54 (65.9) | 0.104 |
| Onset/LKW-to-imaging, h, median (IQR) | 3.0 (1.5-6.7) | 3.2 (1.7-6.1) | 1.9 (1.2-7.9) | 2.1 (1.4-4.3) | 0.070 |
| Baseline NIHSS, median (IQR) | 2 (1-4) | 2 (1-3) | 3 (1-4) | 3.5 (2-4) | <0.001 |
| NIHSS 0, n (%) | 73 (13.7) | 59 (15.1) | 9 (14.8) | 5 (6.1) |  |
| NIHSS 1-2, n (%) | 209 (39.1) | 171 (43.7) | 19 (31.1) | 19 (23.2) |  |
| NIHSS 3-5, n (%) | 252 (47.2) | 161 (41.2) | 33 (54.1) | 58 (70.7) | <0.001 |
| PRISMS-defined nondisabling MIS, n (%) | 284 (53.2) | 230 (58.8) | 31 (50.8) | 23 (28.0) | <0.001 |
| ARAMIS-defined nondisabling MIS, n (%) | 259 (48.5) | 211 (54.0) | 28 (45.9) | 20 (24.4) | <0.001 |
| Oxfordshire: PACI, n (%) | 205 (38.4) | 112 (28.6) | 40 (65.6) | 53 (64.6) |  |
| Oxfordshire: POCI, n (%) | 76 (14.2) | 54 (13.8) | 3 (4.9) | 19 (23.2) |  |
| Oxfordshire: LACI, n (%) | 253 (47.4) | 225 (57.5) | 18 (29.5) | 10 (12.2) | <0.001 |
| TOAST: large-artery atherosclerosis, n (%) | 100 (18.7) | 62 (15.9) | 14 (23.0) | 24 (29.3) |  |
| TOAST: cardioembolism, n (%) | 104 (19.5) | 62 (15.9) | 15 (24.6) | 27 (32.9) |  |
| TOAST: small-vessel occlusion, n (%) | 165 (30.9) | 165 (42.2) | 0 | 0 |  |
| TOAST: undetermined, n (%) | 139 (26.0) | 92 (23.5) | 26 (42.6) | 21 (25.6) |  |
| TOAST: other determined cause, n (%) | 26 (4.9) | 10 (2.6) | 6 (9.8) | 10 (12.2) | <0.001 |
| Tmax >6 s volume, mL, median (IQR) | 0 (0-4) | 0 (0-0) | 13 (5-32) | 25 (13-51) | <0.001 |
| Mismatch volume, mL, median (IQR) | 0 (0-4) | 0 (0-0) | 12 (5-32) | 21 (13-47) | <0.001 |
| Intracranial LVO, n (%) | 53 (9.9) | 0 | 0 | 53 (64.6) | <0.001 |
| Intracranial MeVO, n (%) | 29 (5.4) | 0 | 0 | 29 (35.4) |  |
| Any reperfusion therapy, n (%) | 100 (18.7) | 34 (8.7) | 13 (21.3) | 53 (64.6) | <0.001 |
| IV thrombolysis alone, n (%) | 72 (13.5) | 34 (8.7) | 13 (21.3) | 25 (30.5) |  |
| Endovascular treatment, n (%) | 28 (5.2) | 0 | 0 | 28 (34.1) |  |
| Early neurological deterioration, n (%) | 43 (8.1) | 26 (6.6) | 3 (4.9) | 14 (17.1) | 0.004 |
| 90-day mRS $\geq 3$, n (%) | 94 (17.6) | 52 (13.3) | 21 (34.4) | 21 (25.6) | <0.001 |

Values are mean (SD), median (IQR), or n (%). P values refer to the overall comparison across the 3 phenotypes; for multi-category variables, the P value is shown on the final category row. LACI indicates lacunar infarction; LKW, last known well; LVO, large-vessel occlusion; MeVO, medium-vessel occlusion; MIS, minor ischemic stroke; mRS, modified Rankin Scale; NIHSS, National Institutes of Health Stroke Scale; PACI, partial anterior circulation infarction; PF, perfusion deficit; POCL, posterior circulation infarction; TOAST, Trial of ORG 10172 in Acute Stroke Treatment; VO, vessel occlusion.

Overall, 100 patients (18.7%) received reperfusion therapy, including 72 (13.5%) treated with intravenous thrombolysis alone and 28 (5.2%) undergoing endovascular treatment. The proportion receiving any reperfusion therapy differed markedly across CT phenotypes, from 8.7% in PF-/VO- to 21.3% in PF+/VO- and 64.6% in PF+/VO+ (Table 1).

### Early Neurological Deterioration

END occurred in 43 patients (8.1%). Its frequency differed across multiparametric CT phenotypes, occurring in 6.6% of PF-/VO-, 4.9% of PF+/VO-, and 17.1% of PF+/VO+ patients (P=0.004; Table 2). Vascular risk factors did not differ materially between patients with and without END; baseline NIHSS and classification as nondisabling MIS according to either PRISMS or ARAMIS criteria were also similar between groups.

**Table 2.**
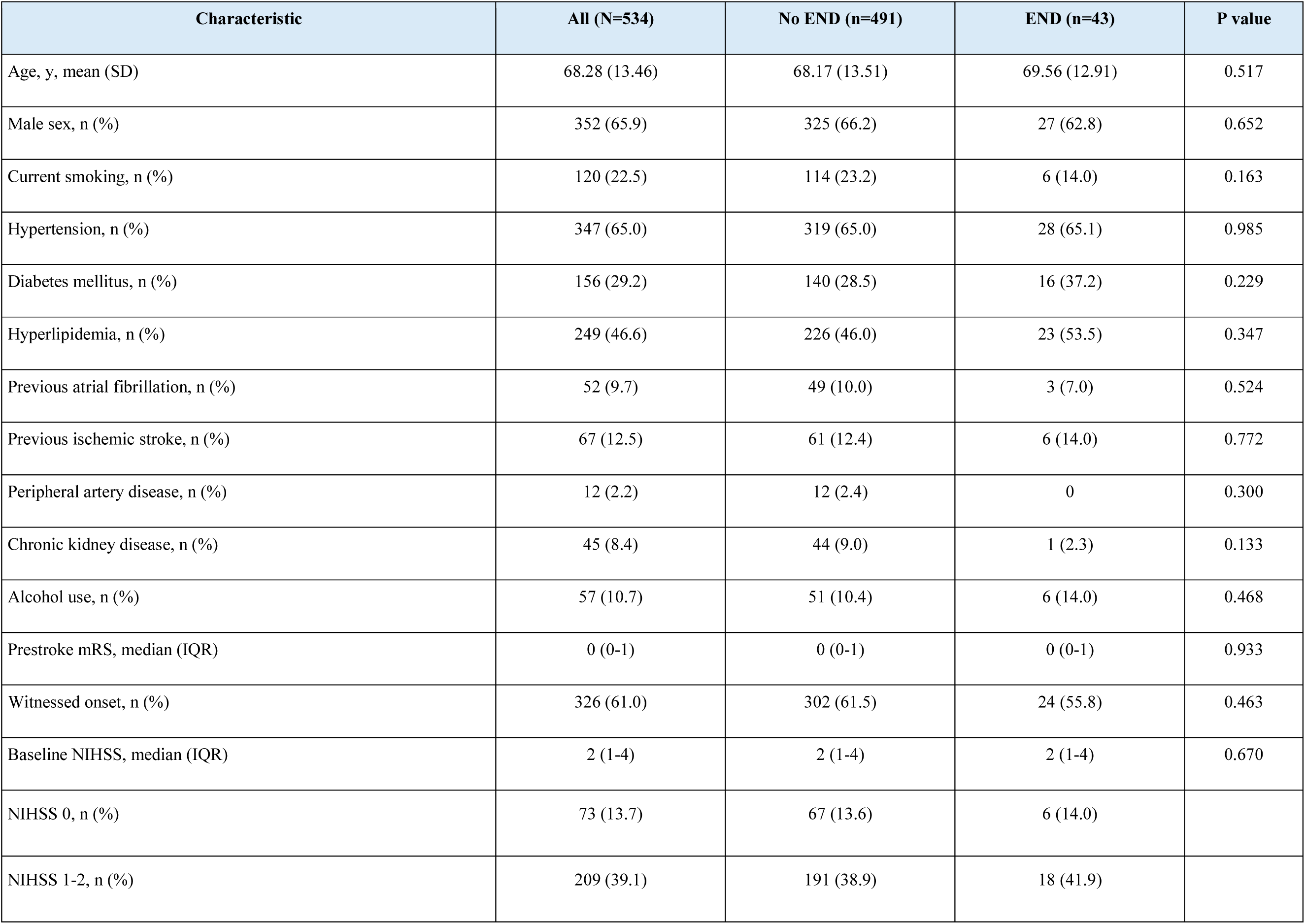
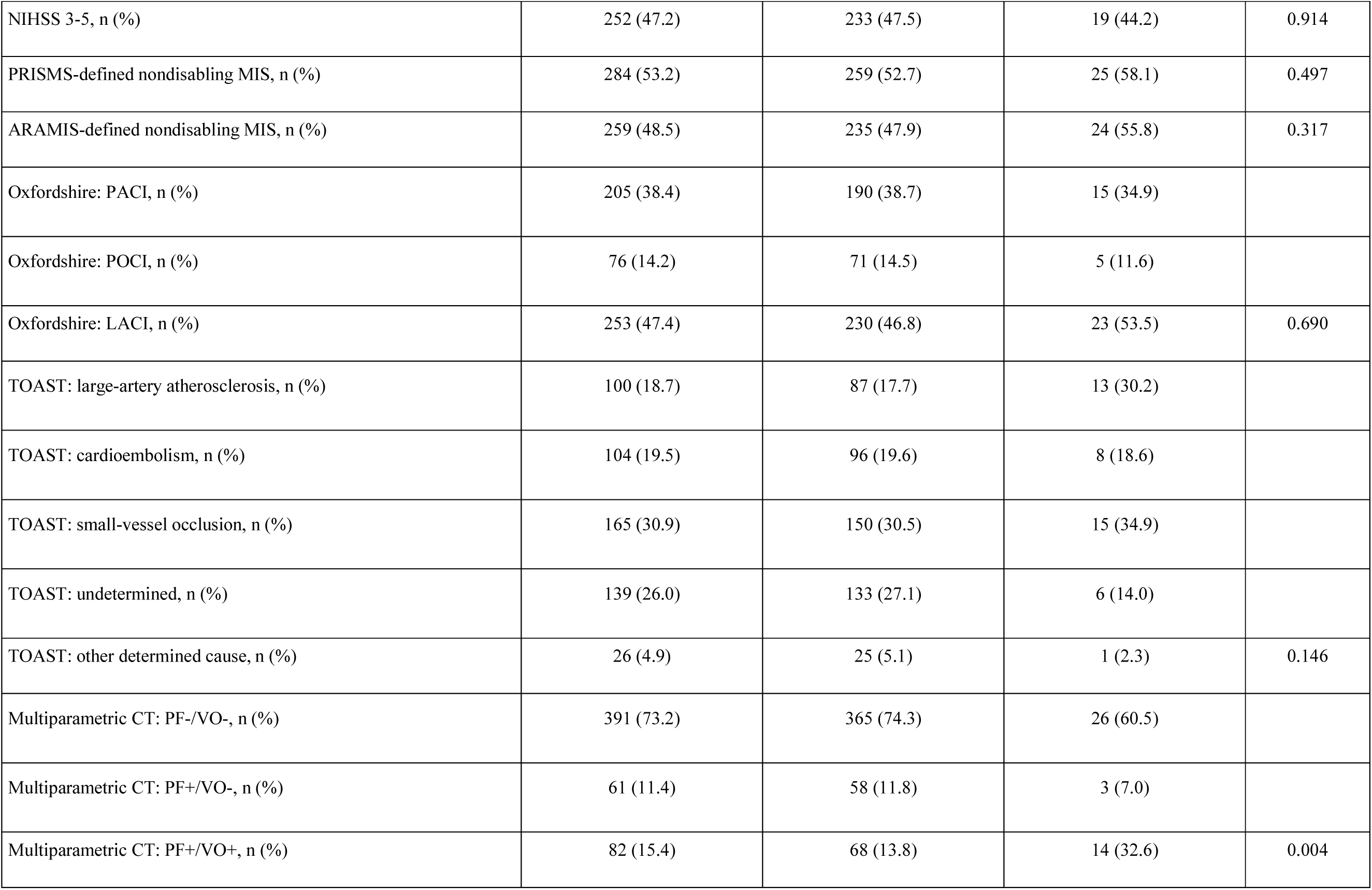
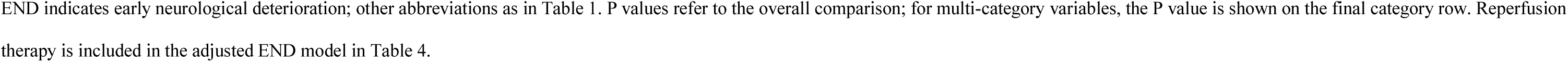
Clinical, Etiologic, and Imaging Characteristics According to Early Neurological Deterioration.

In the multivariable analysis, compared with PF-/VO-, PF+/VO+ was associated with END (aOR, 2.50 [95% CI, 1.03-6.06]; P=0.042), whereas PF+/VO- was not (aOR, 0.71 [95% CI, 0.20-2.46]; P=0.587; Table 4).

### Functional Outcome at 90 Days

The primary outcome, mRS ≥3 at 90 days, occurred in 94 patients (17.6%). Patients with mRS ≥3 were older and more frequently had hypertension, diabetes mellitus, previous atrial fibrillation, and previous ischemic stroke. They also had greater baseline neurological severity, higher prestroke mRS, and were less frequently classified as having nondisabling MIS according to either PRISMS or ARAMIS criteria (Table 3).

**Table 3.** Clinical, Etiologic, Imaging, and Treatment Characteristics According to 90-Day Functional Outcome.

| Characteristic | All (N=534) | mRS <3 (n=440) | mRS ≥3 (n=94) | P value |
| --- | --- | --- | --- | --- |
| Age, y, mean (SD) | 68.28 (13.46) | 66.91 (13.41) | 74.72 (11.77) | <0.001 |
| Male sex, n (%) | 352 (65.9) | 289 (65.7) | 63 (67.0) | 0.804 |
| Current smoking, n (%) | 120 (22.5) | 104 (23.6) | 16 (17.0) | 0.163 |
| Hypertension, n (%) | 347 (65.0) | 272 (61.8) | 75 (79.8) | <0.001 |
| Diabetes mellitus, n (%) | 156 (29.2) | 116 (26.4) | 40 (42.6) | 0.002 |
| Hyperlipidemia, n (%) | 249 (46.6) | 202 (45.9) | 47 (50.0) | 0.470 |
| Previous atrial fibrillation, n (%) | 52 (9.7) | 36 (8.2) | 16 (17.0) | 0.009 |
| Previous ischemic stroke, n (%) | 67 (12.5) | 49 (11.1) | 18 (19.1) | 0.033 |
| Peripheral artery disease, n (%) | 12 (2.2) | 8 (1.8) | 4 (4.3) | 0.148 |
| Chronic kidney disease, n (%) | 45 (8.4) | 36 (8.2) | 9 (9.6) | 0.659 |
| Alcohol use, n (%) | 57 (10.7) | 47 (10.7) | 10 (10.6) | 0.990 |
| Prestroke mRS, median (IQR) | 0 (0-1) | 0 (0-1) | 1 (0-2) | <0.001 |
| Witnessed onset, n (%) | 326 (61.0) | 275 (62.5) | 51 (54.3) | 0.137 |
| NIHSS 0, n (%) | 73 (13.7) | 67 (15.2) | 6 (6.4) |  |
| NIHSS 1-2, n (%) | 209 (39.1) | 188 (42.7) | 21 (22.3) |  |
| NIHSS 3-5, n (%) | 252 (47.2) | 185 (42.0) | 67 (71.3) | <0.001 |
| PRISMS-defined nondisabling MIS, n (%) | 284 (53.2) | 255 (58.0) | 29 (30.9) | <0.001 |
| ARAMIS-defined nondisabling MIS, n (%) | 259 (48.5) | 230 (52.3) | 29 (30.9) | <0.001 |
| Oxfordshire: PACI, n (%) | 205 (38.4) | 164 (37.3) | 41 (43.6) |  |
| Oxfordshire: POCI, n (%) | 76 (14.2) | 62 (14.1) | 14 (14.9) |  |
| Oxfordshire: LACI, n (%) | 253 (47.4) | 214 (48.6) | 39 (41.5) | 0.431 |
| TOAST: large-artery atherosclerosis, n (%) | 100 (18.7) | 76 (17.3) | 24 (25.5) |  |
| TOAST: cardioembolism, n (%) | 104 (19.5) | 82 (18.6) | 22 (23.4) |  |
| TOAST: small-vessel occlusion, n (%) | 165 (30.9) | 144 (32.7) | 21 (22.3) |  |
| TOAST: undetermined, n (%) | 139 (26.0) | 117 (26.6) | 22 (23.4) |  |
| TOAST: other determined cause, n (%) | 26 (4.9) | 21 (4.8) | 5 (5.3) | 0.148 |
| Intracranial vessel status: no VO, n (%) | 452 (84.6) | 379 (86.1) | 73 (77.7) |  |
| Intracranial vessel status: LVO, n (%) | 53 (9.9) | 39 (8.9) | 14 (14.9) |  |
| Intracranial vessel status: MeVO, n (%) | 29 (5.4) | 22 (5.0) | 7 (7.4) | 0.097 |
| Any reperfusion therapy, n (%) | 100 (18.7) | 79 (18.0) | 21 (22.3) | 0.322 |
| Early neurological deterioration, n (%) | 43 (8.1) | 23 (5.2) | 20 (21.3) | <0.001 |
| Multiparametric CT: PF-/VO-, n (%) | 391 (73.2) | 339 (77.0) | 52 (55.3) |  |
| Multiparametric CT: PF+/VO-, n (%) | 61 (11.4) | 40 (9.1) | 21 (22.3) |  |
| Multiparametric CT: PF+/VO+, n (%) | 82 (15.4) | 61 (13.9) | 21 (22.3) | <0.001 |

The primary outcome was unfavorable functional outcome, defined as mRS ≥3 at 90 days. P values refer to the overall comparison; for multi-category variables, the P value is shown on the final category row.

The proportion with mRS ≥3 differed across CT phenotypes: 13.3% in PF-/VO-, 34.4% in PF+/VO-, and 25.6% in PF+/VO+ (P<0.001; Table 3). Receipt of any reperfusion therapy was not associated with the primary outcome in univariable analysis (P=0.322). END was strongly associated with subsequent functional outcome: 46.5% of patients with END had mRS ≥3 compared with 15.1% of those without END (P<0.001), corresponding to an unadjusted OR of 4.90 (95% CI, 2.56-9.37).

In the multivariable analysis, compared with PF-/VO-, PF+/VO- was associated with more than 3-fold higher odds of mRS ≥3 (aOR, 3.36 [95% CI, 1.67-6.73]; P<0.001), whereas PF+/VO+ was not (aOR, 1.58 [95% CI, 0.75-3.34]; P=0.226). Increasing age (aOR per year, 1.043 [95% CI, 1.018-1.068]; P<0.001), diabetes mellitus (aOR, 1.79 [95% CI, 1.05-3.06]; P=0.033), and prestroke mRS 2 versus 0 (aOR, 3.17 [95% CI, 1.63-6.16]; P<0.001) were also independently associated with unfavorable outcome. Any reperfusion therapy was not independently associated with mRS ≥3 (aOR, 0.75 [95% CI, 0.36-1.56]; P=0.438; Table 4).

**Table 4.** Multivariable Logistic Regression Analyses for Early Neurological Deterioration and Unfavorable 90-Day Functional Outcome.

| Variable | END, aOR (95% CI) | P value | mRS $\geq 3$ at 90 d, aOR (95% CI) | P value |
| --- | --- | --- | --- | --- |
| Age, per 1-y increase | 1.008 (0.982-1.034) | 0.541 | 1.043 (1.018-1.068) | <0.001 |
| PF-/VO- | Reference |  | Reference |  |
| PF+/VO- | 0.709 (0.204-2.456) | 0.587 | 3.358 (1.674-6.733) | <0.001 |
| PF+/VO+ | 2.501 (1.032-6.061) | 0.042 | 1.584 (0.753-3.335) | 0.226 |
| Hypertension | - | - | 1.324 (0.715-2.453) | 0.372 |
| Diabetes mellitus | - | - | 1.792 (1.048-3.064) | 0.033 |
| Previous atrial fibrillation | - | - | 0.969 (0.461-2.036) | 0.934 |
| Previous ischemic stroke | - | - | 1.285 (0.636-2.595) | 0.485 |
| NIHSS 0 | Reference |  | Reference |  |
| NIHSS 1-2 | 1.102 (0.409-2.968) | 0.848 | 1.257 (0.446-3.540) | 0.665 |
| NIHSS 3-5 | 0.968 (0.275-3.404) | 0.959 | 2.476 (0.782-7.846) | 0.123 |
| PRISMS-defined nondisabling MIS | 1.675 (0.634-4.423) | 0.298 | 0.534 (0.252-1.132) | 0.102 |
| Prestroke mRS 0 | Reference |  | Reference |  |
| Prestroke mRS 1 | 0.641 (0.257-1.595) | 0.339 | 0.976 (0.512-1.863) | 0.942 |
| Prestroke mRS 2 | 1.375 (0.569-3.323) | 0.479 | 3.165 (1.627-6.159) | <0.001 |
| Any reperfusion therapy | 1.739 (0.714-4.236) | 0.223 | 0.747 (0.358-1.560) | 0.438 |

All covariates were entered simultaneously. The END model included age, CT phenotype, baseline NIHSS category, PRISMS-defined nondisabling MIS, prestroke mRS category, and any reperfusion therapy. The mRS model additionally included hypertension, diabetes mellitus, previous atrial fibrillation, and previous ischemic stroke because these were associated with the outcome in univariable analysis at $P < 0.05$ . PF-/VO-, NIHSS 0, and prestroke mRS 0 were reference categories. aOR indicates adjusted odds ratio; CI, confidence interval.

### Sensitivity Analyses

Replacing PRISMS-defined nondisabling MIS with the ARAMIS definition yielded consistent results (Table S1). PF+/VO+ remained associated with END (aOR, 2.50 [95% CI, 1.03-6.09]; P=0.043), whereas PF+/VO- remained strongly associated with mRS ≥3 (aOR, 3.33 [95% CI, 1.66-6.67]; P<0.001).

Because all 165 patients with lacunar etiology were included in the PF-/VO- group, analyses were repeated after excluding lacunar strokes, leaving 369 patients. In this restricted cohort, END occurred in 4.9% of PF-/VO-, 4.9% of PF+/VO-, and 17.1% of PF+/VO+ patients (P=0.001; Table S2). In adjusted analyses, PF+/VO+ was associated with higher odds of END using either the PRISMS model (aOR, 4.43 [95% CI, 1.53-12.79]; P=0.006) or the ARAMIS model (aOR, 4.47 [95% CI, 1.55-12.86]; P=0.005; Table S3).

The association between CT phenotype and 90-day functional outcome was also preserved after exclusion of lacunar strokes (Tables S4 and S5). PF+/VO- remained independently associated with mRS ≥3 both in the PRISMS model (aOR, 3.71 [95% CI, 1.75-7.85]; P<0.001) and the ARAMIS model (aOR, 3.67 [95% CI, 1.74-7.75]; P<0.001), whereas PF+/VO+ was not independently associated with the primary functional outcome.

## Discussion

In this consecutive cohort of patients with minor ischemic stroke evaluated with multiparametric CT, we identified distinct imaging phenotypes associated with different patterns of early and long-term risk. A perfusion deficit in the absence of a visible intracranial vessel occlusion (PF+/VO-) was independently associated with unfavorable functional outcome at 90 days, whereas the combination of perfusion deficit and vessel occlusion (PF+/VO+) was associated with early neurological deterioration. These associations persisted after adjustment for baseline neurological severity, prestroke disability, and disabling status. Together, these findings suggest that tissue perfusion and vessel status provide complementary, rather than interchangeable, prognostic information in minor ischemic stroke.

These findings may have immediate implications for risk stratification during the initial emergency evaluation of MIS. Multiparametric CT may help identify patients who, despite mild presenting symptoms, remain at substantial risk of deterioration or subsequent disability and who may therefore warrant more individualized monitoring, treatment, or inclusion in future therapeutic trials. To date, randomized trials have generally selected patients primarily on the basis of clinical severity or disabling status rather than on the combined assessment of tissue perfusion and vessel status. PUMICE broadly enrolled patients with NIHSS scores ≤5 within 4.5 hours and compared intravenous prourokinase with standard care, without demonstrating superiority for excellent 90-day outcome.^10^ PRISMS specifically enrolled minor nondisabling stroke and compared alteplase with aspirin but was terminated early and did not demonstrate improved functional outcome with thrombolysis.^8^ ARAMIS used a more structured definition of nondisabling MIS and showed dual antiplatelet therapy to be noninferior to alteplase.^9^ Collectively, these trials have not established superiority of intravenous thrombolysis over contemporary medical therapy when patients are selected predominantly according to NIHSS and clinical disability.

The uncertainty emerging from individual trials is reinforced by a recent meta-analysis restricted to randomized evidence in minor stroke. Across 4 RCTs specifically enrolling 3364 patients with NIHSS ≤5, intravenous thrombolysis did not increase the odds of excellent 90-day functional recovery compared with nonthrombolytic standard care and was associated with lower odds of functional independence and higher odds of symptomatic intracranial hemorrhage and mortality.^24^ Most participants had nondisabling deficits, however, limiting extrapolation to patients with disabling MIS. These findings further suggest that clinical severity alone may be insufficient to identify patients with a favorable risk-benefit profile for reperfusion therapy.

Our findings suggest that multiparametric CT may provide an additional dimension for prognostic stratification and future trial enrichment, including in patients evaluated beyond conventional thrombolysis windows and up to 24 hours from last known well. Particular attention may be warranted for PF+/VO-. Although these patients had no visible intracranial occlusion, approximately one third had an unfavorable 90-day outcome, and PF+/VO- remained strongly associated with mRS ≥3 after multivariable adjustment. Nevertheless, only 21.3% received reperfusion therapy, substantially fewer than the 64.6% of PF+/VO+ patients. This observation should not be interpreted as evidence that PF+/VO- patients were undertreated or that thrombolysis would necessarily improve their outcome. Rather, it suggests that reliance on angiographic findings and clinical severity alone may fail to identify a subgroup with considerable residual risk. Possible mechanisms include distal occlusions below routine CTA resolution, partial or early recanalization with persistent downstream hypoperfusion, collateral dysfunction, or microvascular impairment.

TEMPO-2 represents the randomized trial that most explicitly incorporated acute neuroimaging into minor-stroke selection, enrolling patients within 12 hours who had either an intracranial occlusion or a focal perfusion abnormality. Tenecteplase did not improve functional outcome compared with standard care.^11^ Importantly, however, TEMPO-2 treated vessel occlusion and focal perfusion abnormality as alternative imaging eligibility criteria. Our findings suggest that these features may not be biologically or prognostically interchangeable: PF+/VO- was predominantly associated with later functional disability, whereas PF+/VO+ was associated with END. A recent secondary analysis of TEMPO-2 further showed that successful recanalization among patients with a visible occlusion was associated with better functional recovery and markedly less stroke progression, underscoring the relevance of vascular patency within the imaging-positive minor-stroke population.^25^ Thus, rather than arguing simply for imaging-based selection, our results support a more granular approach integrating tissue perfusion and vessel status.

PF+/VO+ appeared to identify a dynamically unstable phenotype. A persistent arterial occlusion accompanied by downstream hypoperfusion implies tissue whose viability remains dependent on collateral flow. Failure of collateral support has been associated with worsening hypoperfusion and infarct growth, while the extent and severity of perfusion abnormalities strongly predict END in minor stroke with LVO.^3,19,20,26^ Persistent or failed recanalization, distal thrombus migration, arterial reocclusion, and recurrent embolic events may further convert an initially mild deficit into clinically important deterioration.^27,28^ In the MINORCAT-END study of thrombolysed minor stroke, END occurred in 12.1% of patients with LVO versus 3.6% without LVO, and LVO was the strongest predictor of deterioration; END itself was strongly associated with poor 3-month outcome.^29^ The close relationship between END and disability in our cohort is consistent with this observation. Randomized programs designed to address thrombectomy in low-NIHSS LVO, including MOSTE and ENDOLOW, may help clarify the optimal acute strategy in this subgroup; definitive evidence remains awaited.^30,31^

END may not, however, be exclusively hemodynamic. Early reocclusion or recurrent thromboembolic events are additional potential mechanisms, particularly after initial reperfusion.^27,28^ This rationale has prompted evaluation of intensified antithrombotic treatment after thrombolysis. ARTIS, which tested early intravenous aspirin after alteplase, was terminated prematurely because excess symptomatic intracranial hemorrhage outweighed potential benefit.^32^ More recently, EAST randomized patients with minor stroke treated with intravenous thrombolysis to clopidogrel plus aspirin or placebo within 6 hours after thrombolysis. Early dual antiplatelet therapy appeared safe but did not improve excellent 90-day functional outcome and did not reduce END.^33^ Likewise, phase III studies of adjunctive argatroban or eptifibatide after intravenous thrombolysis did not reduce disability.^34,35^ These neutral results argue against routine post-thrombolysis intensification without better biological selection.

At the same time, early recurrence and thrombotic progression remain relevant to the biology of END outside the immediate post-thrombolysis setting. CHANCE^36^ and POINT^37^ established the benefit of early dual antiplatelet therapy for selected minor noncardioembolic stroke or high-risk transient ischemic attack, with much of the benefit occurring early after treatment initiation; INSPIRES^38^ extended evidence to patients treated within 72 hours with presumed atherosclerotic disease. ATAMIS^39^ directly evaluated END as a primary outcome in mild-to-moderate stroke not treated with thrombolysis or thrombectomy and found less neurological deterioration with clopidogrel plus aspirin than with aspirin alone. A prespecified post hoc analysis of ARAMIS further suggested that the relation between antithrombotic strategy and END may depend on vessel status: DAPT was associated with less END than alteplase in patients without LVO, whereas no difference was observed in the small LVO subgroup; the interaction was only marginally significant.^2^ More recently, a target trial emulation comparing intravenous tirofiban with alteplase in 677 patients with minor stroke treated within 4.5 hours found no significant difference in excellent 90-day functional outcome, but tirofiban was associated with substantially less END at 24 hours and fewer bleeding events.^40^ Because this comparison was observational, these findings should be considered hypothesis generating and require randomized confirmation. Collectively, these data support the concept that mechanisms of END, and potentially the optimal preventive strategy, may differ according to the vascular and perfusion phenotype. Our study should therefore be viewed as hypothesis generating: whether PF+/VO+ or PF+/VO- identifies patients with a greater net benefit from reperfusion, DAPT, tirofiban, or other adjunctive strategies requires prospective randomized testing.

Another relevant observation is that the prognostic information provided by multiparametric CT did not depend on how nondisabling stroke was operationalized. Replacing the PRISMS definition with the ARAMIS definition produced essentially unchanged phenotype estimates, and exclusion of lacunar strokes strengthened rather than attenuated the key associations. This is clinically important because the disabling/nondisabling distinction remains context dependent and may not fully capture tissue-level risk. Multiparametric CT therefore appears to provide information that is complementary to, rather than a surrogate for, neurological severity or clinical disability classification.

This study has several strengths. Patients were drawn from a consecutive stroke-code population rather than selected according to vessel occlusion or a specific treatment strategy, allowing evaluation of the spectrum of perfusion-vessel phenotypes encountered in routine practice. Multiparametric CT was systematically incorporated into the emergency pathway; occlusion status and RAPID-derived perfusion data underwent independent neuroradiology review, with abnormalities attributable to artifacts or technical limitations excluded before phenotype assignment; and the 90-day functional assessment was performed blinded to acute imaging results. The consistency of the findings using two definitions of nondisabling stroke and after excluding lacunar etiology further supports their robustness.

Several limitations should also be acknowledged. First, this was a single-center observational study and requires external validation. PF+/VO- and PF+/VO+ were substantially smaller than PF-/VO-, and the number of END events was limited, resulting in wide confidence intervals. Second, reperfusion treatment was not randomized and was strongly influenced by clinical and imaging characteristics; no causal inference regarding treatment efficacy can therefore be made. Third, PF+/VO- is likely mechanistically heterogeneous, and the study cannot determine whether hypoperfusion reflected distal occlusion, early recanalization, collateral dysfunction, microvascular impairment, or another mechanism. Fourth, dichotomizing perfusion and vessel status facilitates clinical interpretation but simplifies continuous hemodynamic information and may be influenced by acquisition, processing, and threshold selection. Finally, excluding the eight patients who died before day 90 and were assigned an mRS score of 6, only 13 patients (2.4% of the total cohort) had functional assessments available only before day 90. Their last available mRS score was carried forward to estimate the 90-day outcome, which may have introduced some outcome misclassification.

In conclusion, multiparametric CT identified prognostically distinct phenotypes among patients with minor ischemic stroke. Perfusion deficit without visible vessel occlusion was independently associated with unfavorable 90-day functional outcome, whereas the combination of perfusion deficit and vessel occlusion identified patients at increased risk of early neurological deterioration. These findings suggest that vessel status and tissue perfusion provide complementary information beyond clinical severity and the disabling/nondisabling classification. Future trials should evaluate whether integrating these imaging phenotypes can improve risk stratification and identify patients most likely to benefit from reperfusion, intensified antithrombotic therapy, or other strategies aimed at preventing neurological deterioration and subsequent disability.

## Authors’ Contributions

Francisco Purroy contributed to study conception and design, investigation, data curation, statistical analysis, funding acquisition, and supervision. Anna Fernández contributed to study design. Laura Pérez-Girona contributed to patient recruitment. Gloria Yhovany Gallego, Albert Freixa-Cruz, Gerard Mauri, and Mikel Vicente contributed to patient recruitment and clinical data collection. Eugènia Saureu and Raquel Mitjana performed the neuroimaging assessment. Ares Peguera, Sara Salvany, Cristina Pereira, and Gloria Arqué contributed to data curation. Alberto Martínez contributed to the investigation. Francisco Purroy, Eugènia Saureu, and Gerard Mauri drafted the manuscript. All authors critically reviewed the manuscript, approved the final version, and agreed to be accountable for the work.

## Declaration of AI Assistance

During manuscript preparation, the authors used ChatGPT (OpenAI) to assist with English-language editing, improve clarity and organization. All AI-assisted content was reviewed and edited by the authors, who take full responsibility for the accuracy, integrity, and originality of the final manuscript.

## Nonstandard Abbreviations and Acronyms

CTP: computed tomography perfusion
DAPT: dual antiplatelet therapy
END: early neurological deterioration
EVT: endovascular treatment
LVO: large-vessel occlusion
MeVO: medium-vessel occlusion
MIS: minor ischemic stroke
PF: perfusion deficit
VO: vessel occlusion

